# Endogenous adenine is a potential driver of the cardiovascular-kidney-metabolic syndrome

**DOI:** 10.1101/2024.08.19.24312277

**Authors:** Ian Tamayo, Hak Joo Lee, M. Imran Aslam, Jian-Jun Liu, Nagarjunachary Ragi, Varsha Karanam, Soumya Maity, Afaf Saliba, Esmeralda Treviño, Huili Zheng, Su Chi Lim, Jan D. Lanzer, Petter Bjornstad, Katherine Tuttle, Kenneth C. Bedi, Kenneth B. Margulies, Vasan Ramachandran, Ahmed Abdel-Latif, Julio Saez-Rodriguez, Ravi Iyengar, Jean C. Bopassa, Kumar Sharma

**Affiliations:** Center for Precision Medicine, University of Texas Health San Antonio.; Division of Cardiology, University of Texas Health San Antonio.; Clinical Research Unit, Khoo Teck Puat Hospital, Singapore; Heidelberg University Hospital, Institute for Computational Biomedicine, Heidelberg, Germany; UW Medicine Diabetes Institute, University of Washington; Department of Medicine, University of Washington, Seattle, WA, USA, Division of Nephrology, Department of Medicine, Kidney Research Institute, University of Washington, Seattle, Washington; Cardiovascular Institute, Perelman School of Medicine, University of Pennsylvania, Philadelphia, Pennsylvania; School of Public Health University of Texas Health San Antonio and University of Texas San Antonio.; Department of Medicine, University of Michigan, Ann Arbor, MI; Department of Pharmacological Sciences and Institute for Systems Biomedicine, Icahn School of Medicine at Mount Sinai, New York, NY; Department of Cellular and Integrative Physiology, University of Texas Health San Antonio, San Antonio, Texas

## Abstract

Mechanisms underlying the cardiovascular-kidney-metabolic (CKM) syndrome are unknown, although key small molecule metabolites may be involved. Bulk and spatial metabolomics identified adenine to be upregulated and specifically enriched in coronary blood vessels in hearts from patients with diabetes and left ventricular hypertrophy. Single nucleus gene expression studies revealed that endothelial methylthioadenosine phosphorylase (MTAP) was increased in human hearts with hypertrophic cardiomyopathy. The urine adenine/creatinine ratio in patients was predictive of incident heart failure with preserved ejection fraction. Heart adenine and MTAP gene expression was increased in a 2-hit mouse model of hypertrophic heart disease and in a model of diastolic dysfunction with diabetes. Inhibition of MTAP blocked adenine accumulation in the heart, restored heart dysfunction in mice with type 2 diabetes and prevented ischemic heart damage in a rat model of myocardial infarction. Mechanistically, adenine-induced impaired mitophagy was reversed by reduction of mTOR. These studies indicate that endogenous adenine is in a causal pathway for heart failure and ischemic heart disease in the context of CKM syndrome.

## Introduction

The cardiovascular-kidney-metabolic (CKM) syndrome is a complex disorder consisting of cardiovascular disease, kidney disease, obesity and Type 2 diabetes. With the growing incidence of obesity worldwide, the CKM has emerged as a major cause of heart dysfunction and heart failure (*1*). Patients with diabetes have a two-fold increased risk of incident heart failure compared to those without diabetes but this risk is often unrecognized in the early stages of disease progression (*2*). Diabetes is a risk factor for heart failure with preserved ejection fraction (HFpEF) and mortality in patients with HFpEF and diabetes is increased by 30-50% when compared to patients with HFpEF without diabetes (*3*). Patients with diabetes without a history of coronary artery disease (CAD) have the same risk of coronary events as those with pre-existing CAD (*4*) and the addition of kidney disease is a strong risk factor for cardiovascular disease (*5*).

Many kidney disease risk factors have been identified for cardiovascular disease in people with diabetes including circulating biomarkers. Specifically, markers of kidney function and inflammation such as cystatin C, kidney injury molecule (KIM1), and soluble tumor necrosis factor (sTNFR) are associated with both kidney and cardiovascular diseases (*6–9*). Kidney disease and heart hypertrophy both involve dysregulation of the mammalian target of rapamycin (mTOR) pathway (*10, 11*). Left ventricular hypertrophy (LVH) is strongly associated with diabetes (*12*) and both LVH and diabetes are risk factors for diastolic dysfunction and HFpEF (*13*). We recently showed that the endogenous small molecule adenine plays a role in kidney failure in patients with diabetes and inhibition of adenine accumulation blocks mTOR activation, diabetic kidney hypertrophy and markers of kidney injury (cystatin C, KIM1 and TNFR1) (*14*). However, the role of dysregulation of small molecule metabolites in the pathogenesis of heart disease in the context of kidney disease and diabetes have not been clearly demonstrated. Due to the overlap of injury markers of cardiovascular disease with diabetes and the link between endogenous metabolites with activation of mTOR in diabetic kidney disease (*15*), the present study examined the potential role of endogenous small molecule metabolites in the development of heart dysfunction with diabetes in people and in pre-clinical models of heart disease using a multi-omic approach.

## Methods

### Human heart procurement and tissue samples

The human cardiac tissues used in this study were obtained from Human Heart Tissue Bank at the University of Pennsylvania. Procurement of human myocardial tissue was performed under protocols and ethical regulations approved by Institutional Review Boards at the University of Pennsylvania (IRB#802781) and the Gift-of-Life Donor Program (Philadelphia, PA) as previously described. Human hearts with LVH and preserved ejection fraction from patients with a diagnosis of diabetes (n=6) were procured at the Hospital of University of Pennsylvania and non-failing hearts were obtained at the time of organ donation from cadaveric donors (n=6). In all cases, hearts were arrested in situ using ice-cold cardioplegia solution and transported to the laboratory on wet ice. Whole hearts and the dissected left ventricle cavities were weighed to determine levels of hypertrophy. Transmural myocardial samples were dissected from the mid LV free wall below the papillary muscle. LV tissues for mass spectrometry were flash frozen in liquid nitrogen within 4 hours of explantation. Contractile parameters, including left ventricle ejection fraction (LVEF), and LV mass were determined by transthoracic echocardiography in patients.

### Animal studies

The db/m and db/db mice were obtained from Jackson Labs. Tissues and blood samples were harvested as approved by IACUC at UTHSA. db/db mice were randomly assigned to treatment with vehicle or methylthio-DADMe-Immucillin-A (MTDIA), methylthioadenosine phosphorylase (MTAP) inhibitor, for a period of 8 weeks from week 8-10 to week 16-18. Heart tissue was split and preserved for both formalin-fixed paraffin-embedded (FFPE) as well as frozen in isopentane with liquid nitrogen immediately after harvest for MALDI-MSI analysis. Frozen tissue was stored in -800C until time of sectioning. Transthoracic echocardiography was performed using a VisualSonics Vevo F2 system with the 57 mhz high frequency transducer (Visual Sonics Inc). Left ventricular ejection fraction, short axis M-mode scans at the mid-ventricular level, Apical 4-chamber to obtain diastolic function measurements using pulsed-wave and tissue Doppler at the level of mitral valve were used in anesthetized mice. Anesthesia was induced by 2-5% isoflurane and transferred to the warmed, temperature-controlled stage for imaging. Isoflurane was reduced to 1.0–1.5% and adjusted to maintain heart rate in the range of 400-500 beats per minute. At the end of the procedures all mice recovered from anesthesia. All parameters were measured at least 3 times, and averages were presented. Phospho-4E-BP1 and 4E-BP1 antibodies were used for immunoblotting. Kidney cortical p21 expression was analyzed by ELISA. mRNA expression of Klotho and GAPDH was measured by RT-qPCR.

### LC-MS untargeted metabolomics of bulk heart tissue

Untargeted LC-MS bulk tissue metabolomic analyses were performed on human left ventricle tissues to identify metabolites that are associated with heart failure. For tissue extraction, 5 mg of heart tissue was homogenized in 25 µL of extraction solution (80% methanol), vortexed for one minute, centrifuged for 8 minutes at 4°C followed by collection of the supernatant which was then diluted 1:10 with extraction solution. LC-MS analysis was performed on Thermo Q Exactive HF-X Orbitrap mass spectrometer in full scan positive ion mode with 100-400 m/z range and interfaced with heated electrospray ionization source (HESI) and coupled to a Thermo Vanquish HPLC system. Acquired Thermo .raw files were converted to vendor neutral .mzML file format using MSConvert software. Full scan LC-MS data was imported to mzMine 3.0 software for *m/z* detection, ion chromatogram building, and chromatographic resolving. Approximately 500 peaks per samples were screened for putative annotation for molecular identification by online database screening using human metabolome database (HMDB) and Kyoto Encyclopedia of Genes and Genomes (KEGG) spectral libraries with 118 molecular IDs being annotated. Peak areas from identified metabolite features were analyzed between healthy control donors and diabetic donors with LVH using Metaboanalyst v6.0 software by volcano plot, PCA, PLS-DA, and heatmap.

### Adenine targeted measurement in urine and bulk heart tissue

For validation and absolute quantification of adenine in heart tissues from the untargeted analysis, adenine was measured in a targeted assay by LC-MS/MS in bulk tissue from human and mouse hearts with the sample preparation method described in the above section. Data was acquired in PRM mode (parallel reaction monitoring). Acquired data was then processed using Xcalibur Quant Browser software and integrated peak area of adenine and the internal standard ratio (A/IS ratio) was compared to a standard curve (0.01-100µM) to calculate the concentration of adenine in heart tissue.

Urine adenine/creatinine concentration were quantified using ZipChip (908 Devices, Boston, MA) capillary electrophoresis platform coupled with mass spectrometry (CE-MS) as described previously. Metabolite separation was achieved with a microfluidic chip that integrates capillary electrophoresis with nano-electrospray ionization through a ZipChip interface. Data acquisition was performed with a Q-Exactive mass spectrometer (Thermo, San Jose, CA) and Thermo Scientific’s software Xcalibur-Quan Browser for data processing. The reportable linear range for urine adenine assay was (0.01-100µM), with a limit of detection at 10 nM and a coefficient of variation (CV) <10% across the reportable linear range.

### MALDI mass spectrometry imaging of heart tissue

Frozen heart tissue from patients or from db/m and db/db mice were sectioned at 7 µm thickness using a Leica CM1950 cryostat onto an indium tin oxide (ITO) glass slide for MSI analysis and adjacent serial sections onto a standard microscope slide for histology. A MALDI matrix, dihydroxybenzoic acid (DHB), was robotically sprayed onto the ITO slide with tissue using the HTX M3+ automated sprayer (40 mg/mL DHB in 50% MeOH, 50 µL/min, 16 passes, 10 psi LN2 sheath gas, 1200 mm/min velocity). MALDI-MSI was performed with a Thermo Q Exactive HF-X orbitrap coupled to a UV-laser MALDI source (Spectroglyph LLC). Data was collected in positive ion mode with 100-1500 m/z range and at a spatial resolution of 20 µm. The raw data files were converted to the common .ibd and .imzML format using ImageInsight software and uploaded to METASPACE, a web-based molecular annotation platform for MADLI-MSI data. Serial frozen heart tissue sections were formalin-fixed (4% formaldehyde) after sectioning and stained with hematoxylin and eosin (H&E) courtesy of the STRL histology core laboratory at UTHSA. Autofluorescence and brightfield (AF-BF) imaging of the H&E-stained slides was performed on the Zeiss Axioscan 7 microscope with 20x objective. High-resolution images were exported to .jpeg format and used for overlay with MSI data to localize metabolites to histology.

### Adenine measurement in human and mouse heart blood vessel

The distribution of adenine in frozen heart tissue sections by MALDI-MSI was analyzed using Bruker SCiLS software. MSI data was uploaded for each sample in SCILS, and pre-MALDI-MSI autofluorescent images were aligned to the total ion current image using a two-point registration strategy. H&E images acquired from adjacent serial tissue sections were overlaid to the AF image to align the MSI data to both optical image types. Blood vessels in hearts were identified and selected from H&E image and their presence was confirmed from the AF image. Only sections which passed tissue quality control parameters, including minimal tissue freezing artifact, were included for coronary blood vessel analysis. A total of 3 blood vessels were identified per heart sample. A region of interest (ROI) was selected encompassing each blood vessel and ion intensity data for adenine (m/z 136.0618 ± 3 ppm) was exported after TIC-normalization for each pixel contained in the ROI. The mean intensity value for adenine was calculated by averaging the intensity of adenine for each pixel. For confirmation of molecular ion identity of adenine, fragmentation of adenine ion (parent m/z = 136.0617) was performed from MALDI experiments using frozen human heart sections (MALDI-MS/MS). Fragment m/z and intensity ratios were compared to human metabolome database (HMDB) spectral library for molecular identity confirmation.

### Single nucleus analysis

Data was downloaded from Broad Institute’s Single Cell Portal (https://singlecell.broadinstitute.org/single_cell) (project ID SCP1303). Cell annotations were manually summarized to main cell lineages. Counts were summed per cell lineage and per patient to psuedobulks. This count matrix was normalized and variance stabilized with edgeR (https://www.ncbi.nlm.nih.gov/pmc/articles/PMC2796818/) and limma (https://academic.oup.com/nar/article/43/7/e47/2414268) R-packages. Differential gene expression analysis was performed using limma, regressing out sex and age. Association between clinical covariates and MTAP expression was assessed with linear models. Enrichment analysis was performed with run ulm() function from the decoupleR (https://academic.oup.com/bioinformaticsadvances/article/2/1/vbac016/6544613) R-package. Rapamycin sensitive genes were acquired from Wikipathways via misgdbr R-package (https://igordot.github.io/msigdbr/authors.html#citation).

### Clinical cohort

The Singapore Study of Macro-Angiopathy and microvascular Reactivity in Type 2 Diabetes (SMART2D) cohort is an ongoing prospective study on vascular and non-vascular complications in Southeast Asian patients with type 2 diabetes. In brief, 2057 outpatients with type 2 diabetes were recruited from a secondary hospital and an adjacent primary care facility between 2011-2014. Participants were followed by reviewing electronic medical records every 2 years and were also invited for in-person research visits in the hospital every 3 years. Data from routine clinical care and research visits were combined to ascertain clinical outcomes. Follow-up was censored on December 31, 2021. We included 661 participants with mild to moderately impaired kidney function (estimated glomerular filtration rate - eGFR 20 to 90 ml/min per 1.73m2) and without severely elevated albuminuria (urine albumin/creatinine ratio (ACR) < 300 mg/g) in the current study. The SMART2D study was approved by the Singapore National Healthcare Ethnic Committee and all participants provided written informed consent.

### Identification of incident heart failure events in SMART2D cohort

Heart failure in SMART2D was diagnosed by cardiologists during routine clinical care and ascertained by reviewing medical records according to the European Society of Cardiology (2016 criteria, i.e. evidence of heart failure by transthoracic echocardiography and N-terminal pro-brain natriuretic peptide (NT-proBNP) > 125 pg/ml) as previously described. Heart failure was subtyped according to left ventricular ejection fraction (LVEF) as heart failure with reduced ejection fraction (HFrEF, LVEF <50%) or preserved ejection fraction (HFpEF, LVEF ≥ 50%).

### Clinical and biochemical variables in SMART2D cohort

Cardiovascular disease (CVD) history, which included nonfatal myocardial infarction, stroke, percutaneous coronary angioplasty, and bypass procedures were self-reported at cohort enrollment and ascertained by reviewing electronic medical records after enrollment. Blood pressure was measured 3 times by a semi-automated blood pressure monitor and the average was used. Total cholesterol and creatinine were measured by enzymatic methods (Roche Cobas Integra 700, Roche Diagnostics, Swiss). HbA1c was measured by a point-of-care immunoassay analyzer (DCA Vantage Analyzer, Siemens, Germany). The eGFR was estimated based on serum creatinine by the 2019 Chronic Kidney Disease Epidemiology Collaboration (CKD-EPI) equation. Urinary albumin was measured by an immunoturbidimetric assay (Roche Cobas c, Roche Diagnostics, Mannheim, Germany) and expressed as an albumin-to-creatinine ratio (ACR, mg/g). Baseline plasma NT-proBNP was measured by proximity extension assay (Olink platform, Uppsala, Sweden).

### Heart tissue RT-PCR gene expression

To measure gene expression in bulk mouse heart tissue, RNA was extracted from the tissue homogenate using the QIAGEN RNeasy Mini Kit (Cat# 74104) according to the manufacturer’s instructions. cDNA was synthesized using the Thermo Fisher Scientific RevertAid Reverse Transcription Kit (Cat# 4374966). The primer sequences for the target genes are displayed in Table S7. The qPCR reaction was performed using the QuantStudio3 platform with SYBR Green PCR Master Mix kit (Thermo Fisher Scientific, Cat# A25742).

### Heart perfusion for myocardial infarct size assessment

All materials used were purchased from Sigma-Aldrich (St. Louis, MO), unless otherwise stated. Male Sprague–Dawley rats (300–350 g) from Charles River were pretreated with the MTAP inhibibtor (MTDIA, 20 mg/kg) or vehicle for four hours and were anesthetized by intraperitoneal injection of ketamine (80 mg-kg−1 i.p.) and xylazine (8 mg-kg−1 i.p.). Heparin (200 Ul/kg) was used to prevent blood coagulation and depth of anaesthesia was monitored by toe pinch reflex and palpebral reflex. The surgical procedures and protocol were similar to those described in our previous study [PMID: 32726689]. Hearts were surgically removed and immediately arrested in cold (4°C) Krebs Henseleit bicarbonate buffer (KH) solution (mM): glucose 11, NaCl 118, KCl 4.7, MgSO4 1.2, KH2PO4 1.2, NaHCO3 25 and CaCl2 3, pH 7.4. The aorta was rapidly cannulated and the heart was retrograde-perfused at a constant rate (12 ml/min) in the Langendorff mode using KH buffer bubbled with 95% O2/5% CO2 at 37 °C. After 30 minutes of equilibration, global normothermic ischemia was induced by stopping KH buffer supply to the aorta for 35 minutes followed by 180 min reperfusion. We preferentially considered good hearts the ones that reached a minimum left ventricular developed pressure (LVDP) of 80 mmHg at the end of the basal perfusion (before ischemia, data not shown). This I/R injury protocol typically results in ∼50% of infarct size [PMID:32726689]. Sham hearts were not subjected to I/R injury but only were perfused for the same duration as the I/R injury protocol.

### Mitophagy assessment

Removal of damaged mitochondria (mitophagy) was investigated using Western blot analysis by determining the impact of the PINK1/Parkin pathway, which is known to be involved in mitophagy via autophagy adaptors (PMID: 23065344), in adenine treated mouse heart. To determine the impact of mitophagy the protein levels of PINK1, Parkin, p-p62, LC3I, LC3II and total ubiquitin was detected in the whole cell lysate from mouse hearts using target-specific antibodies.

### Lipofuscin in human heart

To measure the deposition of lipofuscin in human heart tissue, an analysis of H&E-stained human heart images was performed with QuPath software (v0.4.4). The H&E staining images which passed tissue quality control were included for lipofuscin analysis for the control (n = 4) and diabetic with LVH (n = 4) samples. A pixel classifier was employed to distinguish eosinophilic area, hematoxylin-positive area (nuclei), and lipofuscin. A Random Trees pixel classifier was trained on annotations for these three classes created from H&E staining of human heart tissue (18 total images). The area of lipofuscin-positive regions was calculated as a percent of the entire tissue section area for each image. Lipofuscin measurements for images corresponding to diabetic and non-diabetic donors were compared with two sections from each donor. For correlation of lipofuscin to total heart adenine levels, an average of the lipofuscin measurements was used from the two measurements per donor.

### Statistical analysis

Urine adenine and urine albumin were evaluated as the ratio to urine creatinine (urine adenine/creatinine ratio, UAdCR, and albumin-to-creatinine ACR, respectively), natural log-transformed, and scaled by standard deviation (SD). Clinical and biochemical variables were presented as mean (SD), median (IQR), or percentile where appropriate, and differences across subgroups were compared by one-way ANOVA. We used the Kaplan-Meier method to plot cumulative risk of incident heart failure stratified by UAdCR level in tertile. We employed cause-specific Cox regression to study the association of UAdCR with incident heart failure. The outcome was time to incident heart failure while death was censored. UAdCR was the exposure and modeled as both a continuous (per one SD) and a categorical (tertile) variable. Covariates in the models were selected based on biological plausibility. We adjusted for age, sex (female as reference), ethnicity (Chinese as reference), active smoking, CVD history (yes or no), body mass index (BMI), diabetes duration, HbA1c, mean artery pressure, total cholesterol, baseline eGFR, and urine ACR in model 1. We additionally adjusted plasma NT-proBNP, the established predictor of heart failure in model 2. When the time to HFpEF was analyzed as outcome, incident HFrEF was censored. As sensitivity analysis, we excluded participants who developed renal events (end stage kidney disease or doubling of serum creatinine) before incident heart failure. We performed competing risk regression (Fine and Gray subdistribution) by taking mortality before occurrence of heart failure as a competing risk. The proportional hazards assumption was examined by entering the product of UAdCR * time (year) as a covariate and by Shoenfeld residual. No violation of the PH assumption was identified. We used penalized spline (degree of freedom= 4) to assess the linearity between UAdCR and risk of heart failure. All clinical variables in the fully adjusted model (multivariable model 2) were included in the clinical variable-based models.

## Results

### Adenine accumulates in hearts from patients with diabetes and correlates with left ventricular mass in human hearts

Human heart tissues were processed for untargeted and targeted metabolomics by LC-MS in hearts from non-diabetic individuals (n=6) and hearts from patients with diabetes and LVH (n=6). Patient characteristics are shown in Table S1. Untargeted bulk tissue metabolomics using a small molecule window (m/z from 100-400) identified adenine as the most significantly increased metabolite in diabetic human hearts with LVH (Fig. 1a). An increase in adenine in human hearts from patients with diabetes and LVH was confirmed by targeted metabolomic analysis (Fig. 1b).

**Fig. 1.**
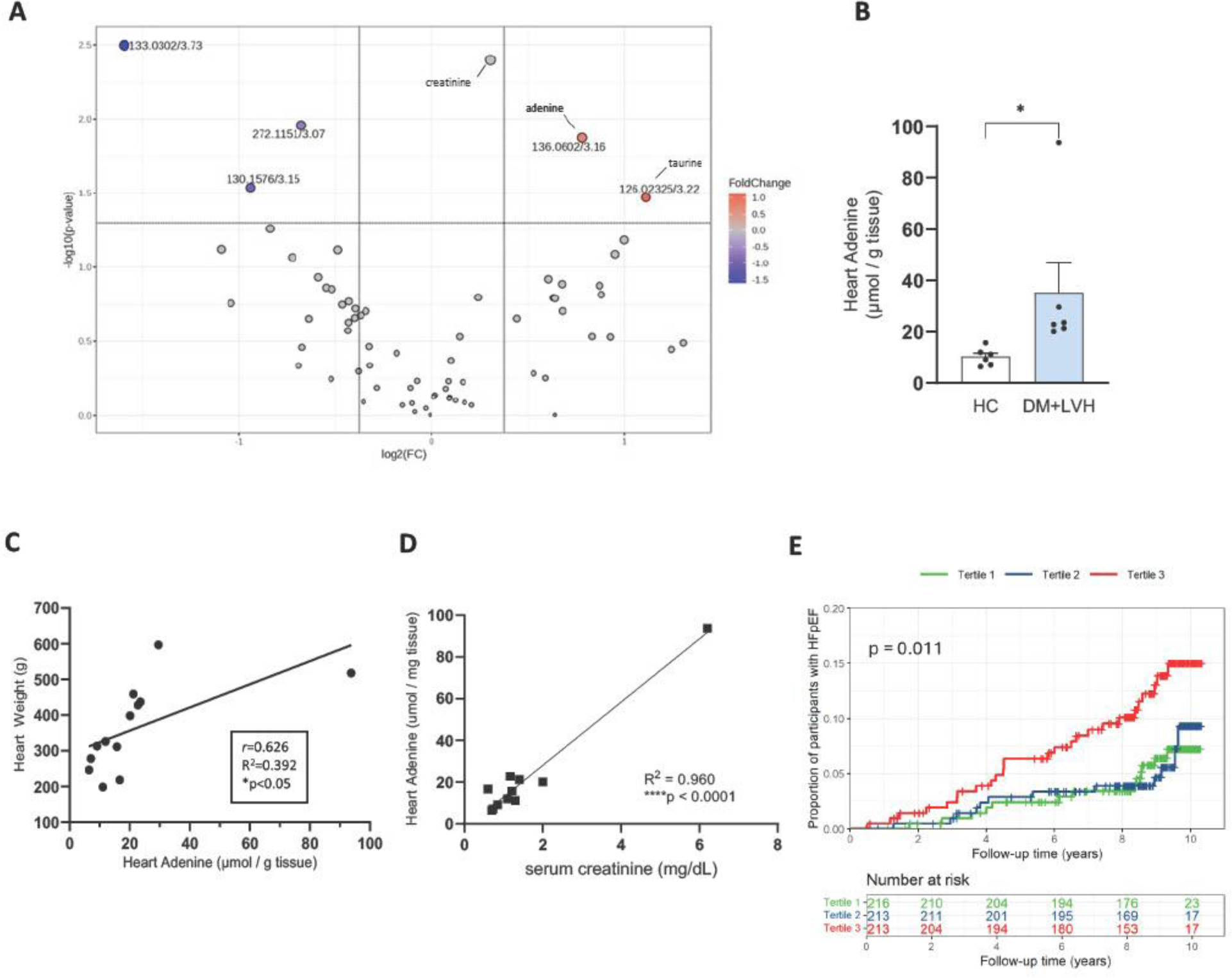
Mass spectrometry reveals increase in adenine in the diabetic patient hearts with LVH. Volcano plot of molecular features identified from an untargeted LC-MS analysis of human donor heart tissues from diabetics with LVH (n=6) and healthy controls (n=6). Fold change cutoff = 1.5 fold, p-value cutoff = 0.05. (**A**). Targeted LC-MS/MS analysis demonstrating increased adenine levels in heart tissue from patients with diabetes and LVH compared to healthy control (**B**). Scatterplot demonstrating human total heart weight is strongly correlated to heart adenine among both diabetics with LVH and healthy control donors (**C**). Scatterplot of heart adenine concentration and serum creatinine levels (Pearson R^2^=0.960, ****p<0.0001) (**D**). Participants from the Singapore Study of Macro-Angiopathy and microvascular Reactivity in Type 2 Diabetes (SMART2D) study (n=653) had UAdCR measurements at the time of enrollment and were followed for 10 years. The participants in the top tertile for UAdCR had the highest risk for incident HFpEF (**E**).

There was a positive correlation of heart adenine with total heart weight (Fig. 1c, r=0.626, p<0.05) and left ventricular mass index (LVMI) across all human hearts evaluated (Fig. S1a, r=0.502, p<0.05). Serum creatinine was also correlated with bulk heart tissue adenine levels, suggesting a link between kidney function and heart adenine (Fig. 1d).

### Urine adenine is associated with risk for HFpEF in patients with type 2 diabetes

To determine if there is a relationship between adenine and heart dysfunction, we analyzed baseline urine adenine/creatinine levels in 653 participants from the Singapore Study of Macro-Angiopathy and microvascular Reactivity in Type 2 Diabetes (SMART2D) study. Patient characteristics are shown in Table S2. 8 patients with prevalent heart failure were excluded from the analysis. As adenine has been identified as a hypertrophic factor in diabetic kidney disease (*14*) and heart disease with diabetes is characterized by cardiac hypertrophy and diastolic dysfunction (*17, 18*), we determined whether UAdCR was associated with development of heart failure with preserved ejection fraction (HFpEF). Participants with a high level of UAdCR had an increased incident risk of HFpEF (Fig. 1e). Cox regression showed one SD increment in UAdCR was associated with a 1.53 (95%CI 1.15-2.03) fold increased risk for HFpEF (Table S3).

Adjustment for cardio-renal risk factors including age, sex, CVD history, SBP, eGFR and urine ACR did not materially alter the association of UAdCR with HFpEF (adjusted HR 1.55 95% CI 1.12-2.15 per 1-SD, multivariable model 1, Table S3). Additional adjustment for the established heart failure biomarker NT-proBNP minimally changed the strength of association (adjusted HR 1.56, 95% CI 1.13-2.16, multivariable model 2). Analyzing UAdCR as a categorical variable gave consistent results (Table S3). Adjusting for competing risk or renal events before HFpEF marginally reduced the hazard ratio in the unadjusted or adjusted models (Table S4, Table S5).

### Spatial metabolomics reveals regional enhancement of adenine in coronary blood vessels with diabetes and snRNAseq reveals enhanced MTAP gene expression in endothelial cells

Spatial metabolomics revealed broad distribution of adenine in human hearts with accumulation in hearts from diabetic donors with LVH (Fig. 2a-c). Multi-modal analysis was performed using autofluorescence and H&E imaging to localize coronary blood vessels (Fig. 1d,e). The spatial mass spectroscopy signal for adenine was registered with the autofluorescence region of the blood vessel (Fig. 1d-g). Via this multi-modal analysis there was a significant increase in adenine in the region of coronary blood vessels in the diabetic hearts compared to the control group (Fig. 1h).

**Fig. 2.**
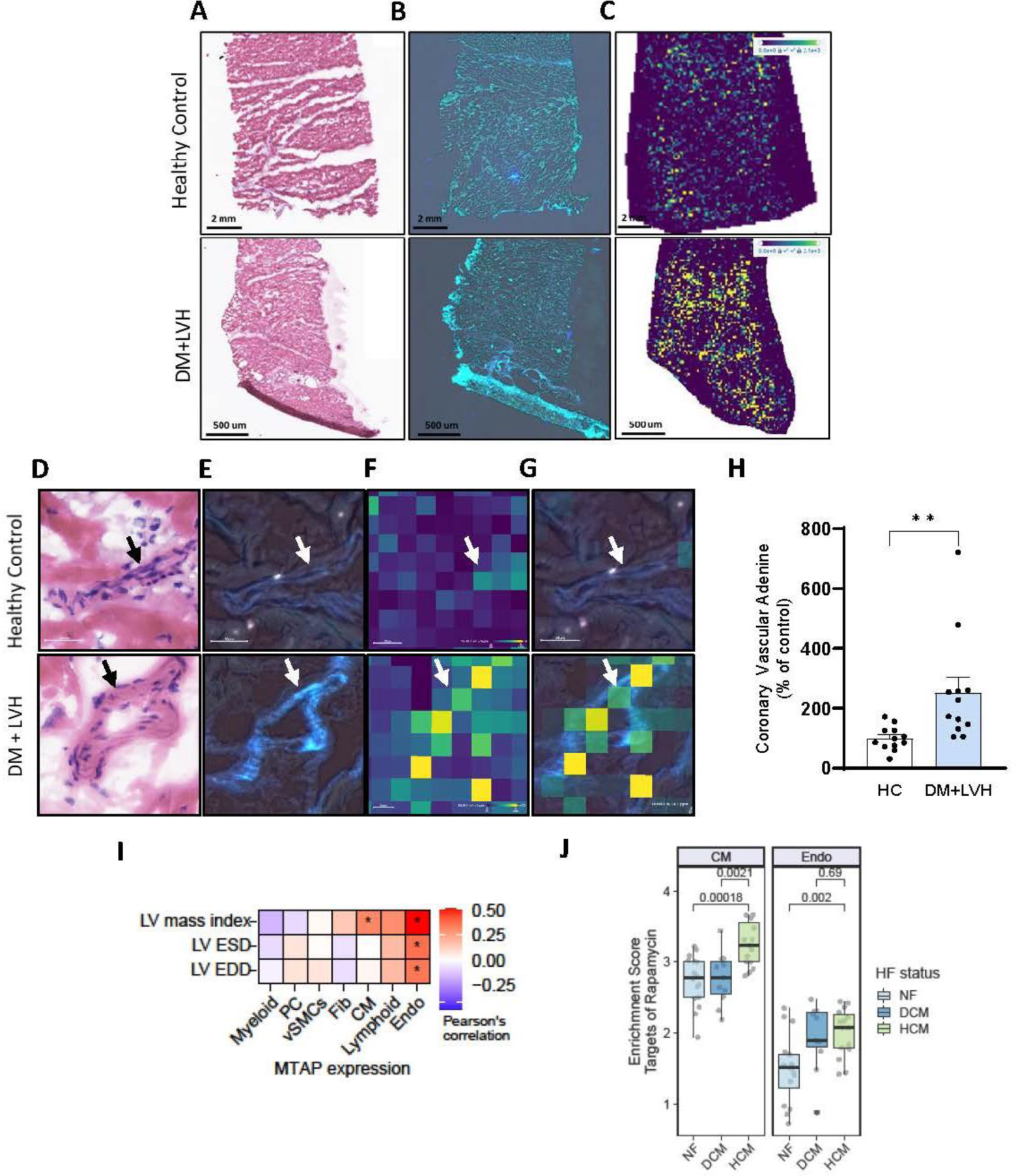
Adenine localizes to cardiac blood vessels in the hearts of patients with diabetes and LVH. H&E staining (**A**), autofluorescent image (**B**), and MALDI-MSI ion image of adenine ([C_5_H_5_N_5_+H]^+^ m/z = 136.0617) (**C**) in a representative healthy control and diabetic patients with LVH. Representative H&E staining image of a blood vessel in the left ventricle from a healthy control patient and a diabetic patient with LVH (**D**). AF imaging of the same blood vessel from adjacent serial section (**E**). MALDI-MSI image of adenine ion ([C_5_H_5_N_5_+H]^+^ m/z = 136.0617) (**F**). Overlay of adenine (MSI) and AF blood vessel image from the same tissue section (**G**). Black/white arrows = blood vessel. Bar graph of the adenine measurement from MSI data for 3 coronary blood vessels from each of control (n=4) and diabetics with LVH (n=4) (**H**) (t-Test, **p-value < 0.01). Pearson correlation coefficients between cell lineage specific MTAP gene expression and cardiac parameters (**I**). Enrichment of rapamycin sensitive genes from WikiPathways in cell lineage specific pseudobulks via univariate linear models (**J**).

We investigated whether upstream and downstream pathways linked to adenine were regulated in human hearts via single nucleus RNAseq analysis from patients with hypertrophic or dilated cardiomyopathy (*16*). The analysis of the data from Chaffin et al (*16*) indicated a positive correlation between endothelial cell-specific MTAP gene expression and LVMI (Fig. 1i). There also was an increased expression of rapamycin sensitive genes in endothelial cells and cardiomyocytes with hypertrophic cardiomyopathy (HCM) (Fig. 1j).

### Enhanced adenine in coronary vessels in mouse models of HFpEF and diabetic heart dysfunction

To address the mechanistic role of endogenous adenine in heart failure, a mouse model of a high fat diet and L-NAME-induced HFpEF (*19*) was used. Heart tissue sections from the left ventricle of HFpEF mouse hearts demonstrated an accumulation of vascular adenine by MALDI-MSI (Fig. 3a-f). *Mtap* gene expression was increased in the left ventricle tissue of HFpEF mice (Fig. 3g). There was a lower relative gene expression of transcription factor A, mitochondrial (*Tfam)* and vascular endothelial growth factor A (*Vegfa)* in the left ventricle tissue of HFpEF mice compared to control (Fig. 3h,i).

**Fig. 3.**
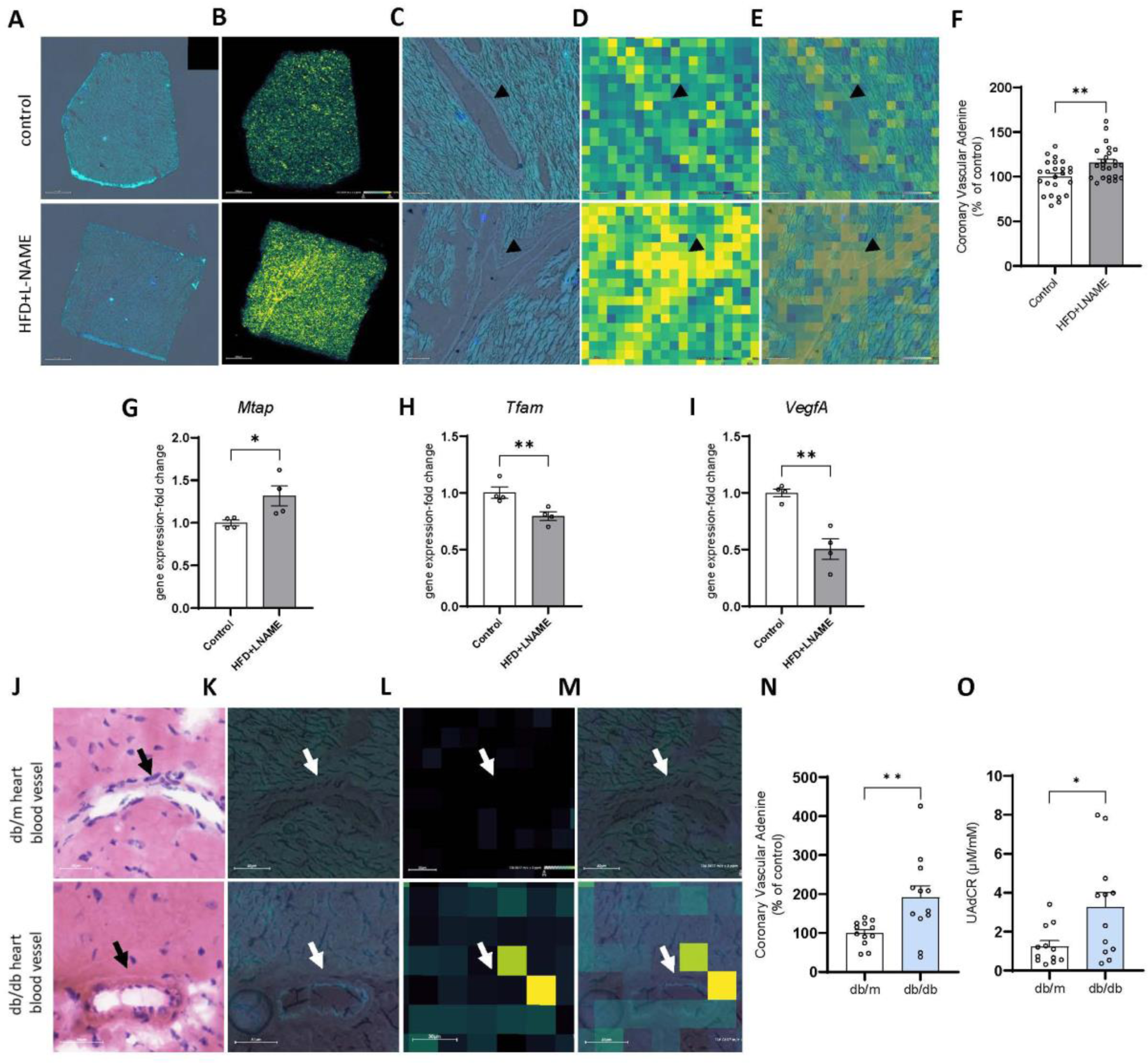
Mass spectrometry reveals increase in heart adenine in mouse models of HFpEF and diabetes. AF images (**A**) and MSI images of adenine ([C_5_H_5_N_5_+Cl^-^]^-^ m/z = 170.0239) in mice treated with HFD + L-NAME and control mice (**A-F**). MALDI-MSI image of adenine ion ([C_5_H_5_N_5_+Cl^-^]^-^ m/z = 170.0239) from the same tissue section (**B**). AF imaging of a coronary blood vessel from HFD + L-NAME-treated and control mice. Black arrows = blood vessel (**C**). Overlay of adenine (from MSI) and AF blood vessel image from the same tissue section (**D** and **E**). Measurement of adenine from MSI data for 3 coronary blood vessels from control (n=4) and HFD + L-NAME treated mouse hearts (n=4) (**F**). Mice treated with HFD + L-NAME show increased *Mtap* gene expression (**G**) and decreased *Tfam* (**H**) and *VegfA* (**I**) gene expression (mean + SE, *p<0.05, **p<0.01, ***p<0.001 by t-test). H&E staining image of a blood vessel in the left ventricle from a db/m and db/db mouse (**J**). Autofluorescence imaging of the same blood vessel from post-MALDI H&E staining (**K**). MALDI-MSI image of adenine ion (m/z = 136.0617) (**L**). Overlay of adenine mass spectrometry image and AF image of blood vessel. Black/white arrows = coronary blood vessel (**M**). Localization of adenine to 4 coronary blood vessels from db/m (n=3) and db/db mouse heart (n=3) (mean + SE) (**N**). Urine adenine/creatinine ratio is increased in db/db mice at 8-10 weeks of age (n=12/group, *p<0.05 (**O**).

To investigate the role of adenine in the context of diabetic heart dysfunction, the spontaneous db/db mouse model was evaluated as cardiac dysfunction has been well described in this model of unabated appetite leading to obesity and diabetes without genetic, chemical or mechanical manipulation (*20, 21*). Spatial metabolomic analysis performed in control and diabetic hearts revealed increased levels of adenine in coronary blood vessels in the db/db mouse (Fig. 3j-n), corresponding to what was observed in both the HFpEF mouse model and hearts from human donors with diabetes and LVH. Interestingly, the db/db mice have increased UAdCR (Fig. 3o). There was a strong positive correlation of bulk heart adenine concentration with both left ventricular mass and total heart weight in the normal and diabetic mice (Fig. 4g) as was observed in the human hearts.

**Fig. 4.**
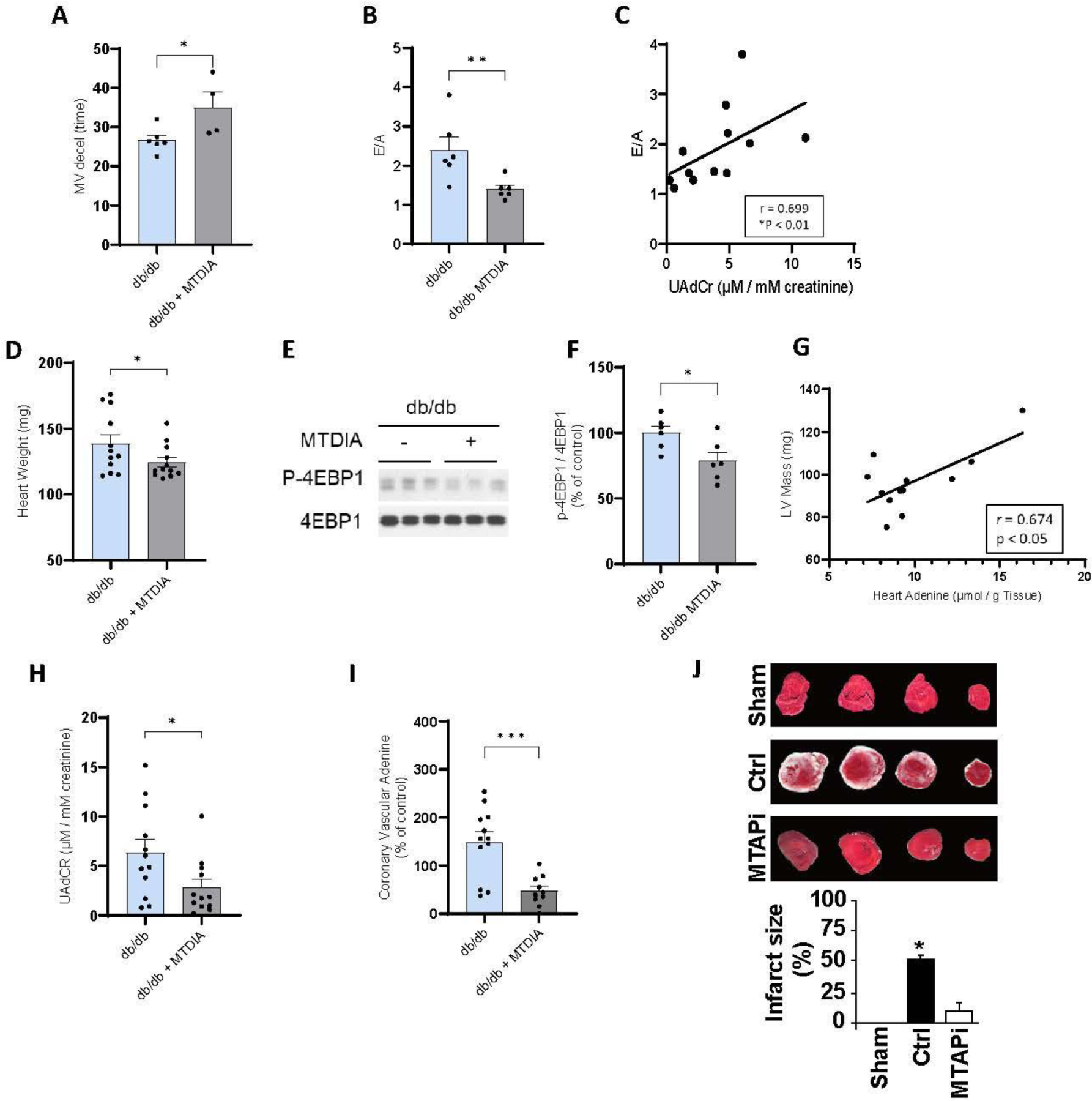
Inhibition of adenine synthesis with MTDIA restores diabetic cardiac dysfunction and reduces heart mTOR activity in diabetic mice. Mitral valve deceleration time was increased in MTDIA-treated db/db mice (**A**) (n=4-6 per group). Echo E/A ratio was reduced in MTDIA-treated db/db mice (**B**). The urine AdCR was correlated with E/A ratio using Spearman correlation (*r* = 0.699) (**C**). Heart adenine was correlated with E/A ratio for all db/db mice with Spearman correlation (n=6 per group) *(r* = 0.6154) (**D**). Western blot of db/db and MTDIA-treated db/db mouse heart measuring p-4EBP1 and 4EBP1 (**E**) demonstrating MTDIA-treatment of db/db mouse significantly reduced mTORC1 activity as measured by p-4EBP1 (n=6 per group) (**F**). Scatterplot of db/db and MTDIA-treated db/db mouse LV mass against heart adenine concentration with Pearson correlation (n=6 per group) (*r* = 0.674, *p < 0.05) (**G**). Bar graph demonstrating decreased urinary adenine to creatinine ratio among MTDIA-treated db/db mice (n=12 per group) (**H**). MSI analysis of adenine in 4 coronary blood vessels from db/db mouse heart and MTDIA-treated db/db mouse heart (n=3 per group) (**I**). Inhibition of MTAP (MTAPi) reduced the infarct size % in a rat model of myocardial infarction (**J**). (mean + SE, *p<0.05, **p<0.01, ***p<0.001 by t-test).

### The role of adenine in diabetic heart dysfunction and myocardial infarction

To determine if there is a causal relationship between adenine and diabetic heart dysfunction, db/db mice were treated with an oral inhibitor of the MTAP enzyme (MTDIA), which blocks the conversion of 5’-methylthioadenosine to adenine. There were no changes in body weight or blood glucose during the study in the db/db mice treated with the MTAP inhibitor (Fig. S3a,b). Heart function was measured by echocardiography in the diabetic mice. The MV deceleration time was reduced in db/db mice (indicating poor diastolic function) and improved in db/db mice treated with the MTAP inhibitor (26.7 ± 3.1 versus 34.8 ± 6.7, p<0.05) (Fig. 4a). The E/A ratio, another echocardiographic marker of diastolic dysfunction, was reduced by the MTAP inhibitor in db/db mice (Fig. 4b). Interestingly, the E/A ratio was positively correlated with the UAdCR (Fig. 4c). MTAP inhibition reduced overall heart weight in db/db mice (Fig. 4d) and prevented activation of mTOR demonstrated by reduced phosphorylation of 4E-BP1 in the db/db mouse heart (Fig. 4e,f). Treatment with MTAP inhibitor reduced overall heart levels of adenine in the db/db mouse heart (Fig. S2a,b), UAdCR (Fig. 4h), and coronary blood vessel adenine (Fig. 4i). There was an overall correlation of UAdCR with heart adenine levels (Fig. S2c) as well as a correlation with both overall weight and LV mass with heart adenine levels (Fig. S2d,e).

As the spatial metabolomic data found localization of adenine in coronary blood vessels, we speculated that adenine accumulation could also play a role in coronary ischemia. To test this question, MTDIA was administered to rats prior to ex-vivo ischemia-reperfusion (I/R). Myocardial infarct size was significantly increased in the control group after I/R but myocardium was preserved in MTDIA-treated animals (52 ± 5 in control hearts versus 11 ± 2 in MTDIA-treated hearts, Fig. 4j). These data demonstrate that the administration of only one bolus of MTAP inhibitor before I/R insult is sufficient to protect the myocardium against I/R injury.

### Effect of elevation of endogenous adenine on cardiac cells

Mitophagy and decreased bioenergetics have been previously linked to heart failure in humans (*22*). We observed an increased number of damaged mitochondria associated with engulfed mitochondria in diabetic human hearts (Fig. 5b,d) which also demonstrated an increase in cardiomyocyte size (hypertrophy) (Fig. 5a,c) correlating with an accumulation of adenine in the heart tissue (Fig. 1c,d). We therefore considered a defect of the removal of damaged mitochondria, known as mitophagy, to play a role in the damaging effect of adenine in the diabetic heart. The mitophagy pathway is characterized by the PTEN-induced putative kinase 1 (PINK1)/Parkin-dependent pathway (*23*). In mitophagy *via* autophagy, the p62 protein binds to ubiquitinated proteins through its ubiquitin-associated domain (*24*) and to LC3 *via* its LC3-interacting region (*25*). LC3I and LC3II are known to interact with p62 to tether mitochondria to the autophagosome (*26*). We, therefore, studied the role of mitophagy in the accumulation of damaged mitochondria in hearts from mice treated with adenine. Administration of adenine in wild-type mice reduced Beclin1, Pink1, LC3I, LC3II, p62 protein levels. In addition, mitochondrial protein ubiquitination was reduced and cytosolic Parkin protein levels were increased (Fig. 5e,f), indicating that adenine impairs cardiac mitophagy. Supplementation with the mTORC1 inhibitor (rapamycin) (*27, 28*) prevented adenine inhibition of mitophagy (Fig. 5e,f). Failure of mitochondria removal and the consequent oxidation of mitochondrial protein contribute to lipofuscin formation (*29*) and lipofuscin was found to correlate with heart adenine concentration in human hearts (R^2^ =0.453, p = 0.047) (Fig. S1b-e).

**Fig. 5.**
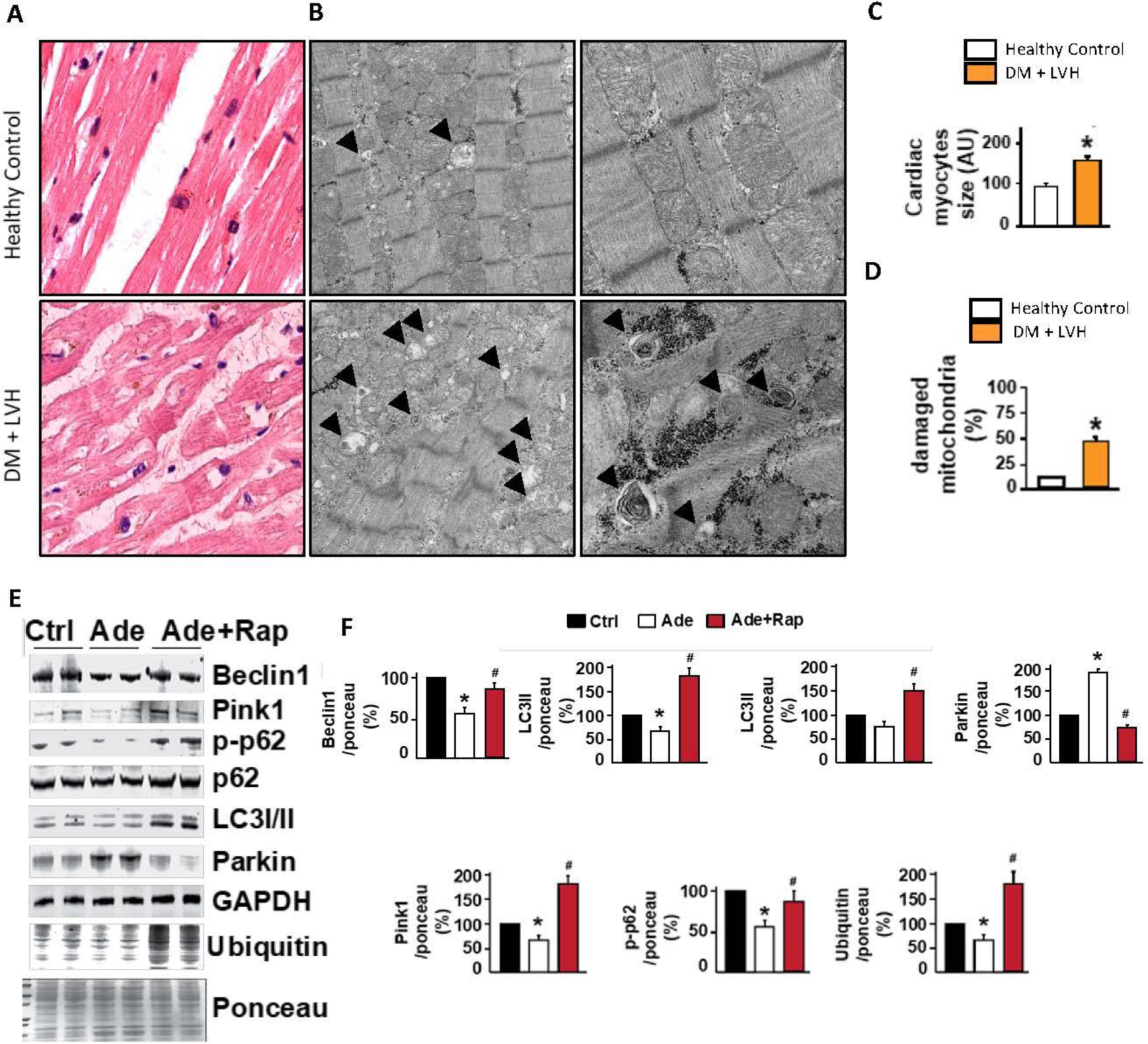
Adenine alters mitophagy in wild-type mice which is reversed with inhibition of mTOR. H&E staining of human hearts from healthy donors and those with diabetes and LVH (**A**). Electron microscopy images of the same cohort. Black arrows point to damaged mitochondria associated with mitochondrial autophagy (**B**). Quantification of cardiomyocyte size from H&E (**C**). Frequency of damaged mitochondria in healthy control and diabetic donors with LVH (**D**). Western blot of whole cell lysate from adenine-treated wild-type mouse heart tissue (**E**) demonstrating that adenine treatment decreased the protein levels of Beclin1, LC3I, LC3II, p-p62, and mitochondrial protein ubiquitination, and increased Parkin expression versus control (**F**). Rapamycin blocked the adenine-induced effects on mitophagy (**E** and **F**). Values are expressed as mean ± SEM; *P < 0.05 control versus control (Ctrl) group, and ^#^P < 0.05 Rapamycin versus Adenine group (n = 3/group).

## Discussion

The basis for the CKM syndrome and HFpEF has remained an enigma. In the present study, we identify a potential explanation for the development of diabetic HFpEF via accumulation of the nucleoside metabolite adenine in the diabetic heart. Adenine was found to accumulate in the hearts of patients with diabetes and correlated with the degree of kidney function. Adenine was specifically increased in coronary blood vessels in humans and mice with diabetes and MTAP gene expression was increased in endothelial cells in human hearts with hypertrophy. Increased UAdCR levels identified patients with diabetes at increased risk for incident HFpEF. Endogenous adenine appears to be in the causative pathway as inhibition of adenine production led to the normalization of the features of diabetic cardiac dysfunction in a diabetic mouse model and inhibition of adenine reduced myocardial infarct size in a rat model.

HFpEF with diabetes is defined as a subtype of heart failure in which there is insufficient cardiac output for the metabolic needs of the individual (*30*). Current hypotheses causing diabetic heart dysfunction include insulin resistance, endothelial dysfunction, cardiac hypertrophy, cardiac fibrosis, cardiac lipotoxicity, advanced glycation end products, matrix remodeling and bioenergetic impairment. Disturbed cardiac metabolism is central to diabetic cardiomyopathy with inefficient substrate utilization, an inability to increase glucose metabolism, and dependence on fatty acid oxidation within the diabetic heart resulting in mitochondrial uncoupling. Despite intensive investigation, the basis for why some patients with T2D develop heart dysfunction is unknown (*30, 31*).

Spatial metabolomics on human heart tissue supports a key role for adenine, as imaging mass spectrometry demonstrated an overall accumulation of adenine in hearts of patients with diabetes, LVH and kidney dysfunction. A functional role for adenine to stimulate heart thickness was suggested as there was a robust correlation of heart adenine content with left ventricular mass and overall heart weight in humans. There was a similar strong correlation of left ventricular mass with heart adenine in mice with diabetes and reduction of adenine in mice with diabetes led to reduction of heart weight. Multiple cell types could contribute to production of adenine due to imbalance in polyamine metabolism, reduction in AMPK activation and enhanced mTORC1 leading to reduced mitophagy. Cardiomyocytes (*32*), vascular smooth muscle cells (*33*), and endothelial cells (*34*) have all been documented to have polyamine pathways.

The causative role of endogenous adenine to diabetic heart dysfunction was tested in a mouse model of type 2 diabetes with diastolic dysfunction (*21*). The db/db mouse has a mutation in the leptin receptor and develops obesity, insulin resistance, and hyperglycemia due to unabated feeding and lack of satiety. The mice develop cardiac dysfunction similar to human diabetic cardiomyopathy (*35*). Inhibition of adenine accumulation via a specific inhibitor of MTAP attenuated features of diastolic dysfunction in the diabetic mice, without affecting blood glucose or body weight. The basis for how adenine contributes to heart dysfunction may be due to impaired mitophagy via mTORC1 as MTAP inhibition led to reduction of mTOR activity in the diabetic heart and adenine-induced impaired mitophagy was blocked by rapamycin. It is well established that in many organ systems mTORC1 activity increases translational capacity, lipid biosynthesis and mitochondrial biogenesis that together can contribute to hypertrophy of cardiomyocytes (*36*) therefore suggesting that endogenous adenine may be a driver of cardiac hypertrophy. There was also a correlation of heart adenine with lipofuscin accumulation suggesting that adenine may contribute to cardiomyocyte senescence and disturbed autophagy. In addition to the role of adenine in cardiac hypertrophy, there is also a role for adenine in coronary vascular function. Spatial metabolomics localized adenine to the coronary vessels and coronary adenine levels were increased in humans and mice with diabetes. A functional role for vascular adenine was demonstrated in a model of acute myocardial infarction as inhibition of adenine production led to protection against ischemic induced myocardial infarct size.

There are several limitations of our study. The urine adenine/creatinine levels were performed in an Asian population (Asian Indians, Chinese and Malays). Validation of the urine adenine studies in other ethnic populations with T2D will assess whether our findings may be generalized to other populations. The reduction of adenine production with the MTAP inhibitor was found to be beneficial in mice and rats but interventional studies in humans have not yet begun. It is of note that we recently found SGLT2 inhibition (empagliflozin) lowered UAdCR levels in patients with diabetes (*14*) and SGLT2 inhibition is well characterized to be heart and kidney protective. Therefore, part of the benefits of SGLT2 inhibition in the CKM syndrome could be via the reduction of heart and kidney adenine levels.

In conclusion, we found the small molecule endogenous metabolite adenine colocalizes with coronary arteries in human hearts with diabetes and that UAdCR predicted the risk of HFpEF in patients with T2D and impaired kidney function. To our knowledge, this is the first description of a metabolite biomarker that predicts HFpEF in patients with diabetes. Our interventional data indicate a causative role for adenine in diabetic cardiomyopathy and ischemic heart disease. Therefore, endogenous adenine production may be a potential key metabolite linking metabolic and kidney dysfunction with cardiovascular disease.

## Data Availability

The code for the QuPath pixel classifier can be found at https://github.com/iantamayo/QuPath-Pixel-Classifier-Heart-H-E-Lipofuscin-Detection. Code for Single nuc analysis can be found at https://github.com/saezlab/adenin_in_hf.

https://github.com/iantamayo/QuPath-Pixel-Classifier-Heart-H-E-Lipofuscin-Detection

## Acknowledgments

We would like to thank the Gift of Life Donor Program of Philadelphia for enabling the procurement of human hearts from deceased organ donors.

## Funding

National Institutes of Health grant UO1DK114920 (K.S.)

Veteran’s Affairs Merit grant I01BX001340 (K.S.)

KTPH STAR Grant 20201 (J.J.L)

KTPH STAR Grant 23201 (J.J.L)

Singapore National Medical Research Council Grant MOH-000066 (S.C.L)

Singapore National Medical Research Council Grant MOH-000714-01 (S.C.L)

Singapore National Medical Research Council Grant MOH-001327-02 (S.C.L)

Federal Ministry of Education and Research Germany (BMBF) 01EJ2201B Curefib (J.D.L.)

National Institutes of Health grant R01MD014712 (K.R.T.)

National Institutes of Health grant U2CDK114886 (K.R.T.)

National Institutes of Health grant UL1TR002319 (K.R.T.)

National Institutes of Health grant U54DK083912 (K.R.T.)

National Institutes of Health grant U01DK100846 (K.R.T.)

National Institutes of Health grant OT2HL161847 (K.R.T.)

National Institutes of Health grant UM1AI109568 (K.R.T.)

Center for Disease Control project number 75D301-21-P-12254 (K.R.T)

National Institutes of Health grant R01HL149891 (K.B.M)

National Institutes of Health grant R01HL105993 (K.B.M)

National Institutes of Health grant R01 GM137056 (RI)

## Author contributions

All authors played a significant role in at least one of these areas: project conceptualization, experimental design, data analysis and interpretation, and preparation of this paper. K.S. is Director of the Center for Precision Medicine (CPM) at UT Health San Antonio and supervised all aspects of the study. J.J.L, H.Z., S.C.L contributed data collection and analysis for the Singapore SMART2D clinical study. K.C.B., K.B.M. contributed human heart tissue and clinical measurements from heart donors. I.T., N.R., S.M., A.S., contributed data collection and analysis of human and mouse heart tissues. H.J.L, I.T. contributed data collection and analysis of mouse tissues. K.S., I.T, H.J.L., R.I., P.B., K.R.T., V.R., J.C.B. made significant contributions to the writing of this paper. J.C.B., V.K., H.J.L., I.A. performed echocardiogram data collection and interpretation in the mouse model data.

## Competing interests

Dr. Margulies holds research grants from Amgen and serves as a scientific consultant/advisory board member for Bristol Myers Squibb and Amgen. Dr. Sharma serves on the data safety board for Cara Therapeutics and holds equity in SygnaMap. All other authors declare that they have no competing interests. Dr. Tuttle has received investigator-initiated grant support (to Providence Inland Northwest Health) from Travere and Bayer outside of the submitted work; consultancy fees from AstraZeneca, Boehringer Ingelheim, Eli Lilly and Company, Novo Nordisk and Travere; speaker fees from AstraZeneca, Eli Lilly, and Novo Nordisk. Dr. Julia Saez-Rodriguez reports funding from GSK, Pfizer and Sanofi & fees/honoraria from Travere Therapeutics, Stadapharm, Astex, Owkin, Pfizer and Grunenthal.

## Notes

### Author Declarations

Procurement of human myocardial tissue was performed under protocols and ethical regulations approved by Institutional Review Boards at the University of Pennsylvania (IRB#802781) and the Gift-of-Life Donor Program (Philadelphia, PA).

## References and Notes

1. L. Athithan, G. S. Gulsin, G. P. McCann, E. Levelt, Diabetic cardiomyopathy: Pathophysiology, theories and evidence to date. World J Diabetes 10, 490–510 (2019).

2. M. Packer, Heart Failure: The Most Important, Preventable, and Treatable Cardiovascular Complication of Type 2 Diabetes. Diabetes Care 41, 11–13 (2018).

3. J. A. Shaw, M. E. Cooper, Contemporary Management of Heart Failure in Patients With Diabetes. Diabetes Care 43, 2895–2903 (2020).

4. J. Cui, Y. Liu, Y. Li, F. Xu, Y. Liu, Type 2 Diabetes and Myocardial Infarction: Recent Clinical Evidence and Perspective. Front Cardiovasc Med 8, 644189 (2021).

5. A. S. Go, G. M. Chertow, D. Fan, C. E. McCulloch, C. Y. Hsu, Chronic kidney disease and the risks of death, cardiovascular events, and hospitalization. N Engl J Med 351, 1296–1305 (2004).

6. J. S. Lees et al., Assessment of Cystatin C Level for Risk Stratification in Adults With Chronic Kidney Disease. JAMA Netw Open 5, e2238300 (2022).

7. I. H. de Boer et al., Cystatin C, albuminuria, and mortality among older adults with diabetes. Diabetes Care 32, 1833–1838 (2009).

8. J. A. Regan et al., Protein biomarkers of cardiac remodeling and inflammation associated with HFpEF and incident events. Sci Rep 12, 20072 (2022).

9. B. N. Putko et al., Circulating levels of tumor necrosis factor-alpha receptor 2 are increased in heart failure with preserved ejection fraction relative to heart failure with reduced ejection fraction: evidence for a divergence in pathophysiology. PLoS One 9, e99495 (2014).

10. M. Godel et al., Role of mTOR in podocyte function and diabetic nephropathy in humans and mice. J Clin Invest 121, 2197–2209 (2011).

11. X. Wu et al., Genetic and pharmacological inhibition of Rheb1-mTORC1 signaling exerts cardioprotection against adverse cardiac remodeling in mice. Am J Pathol 182, 2005–2014 (2013).

12. J. Lv et al., Relationship Between Left Ventricular Hypertrophy and Diabetes Is Likely Bidirectional: A Temporality Analysis. J Am Heart Assoc 12, e028219 (2023).

13. A. A. Oktay, T. K. Paul, C. A. Koch, C. J. Lavie, in Endotext, K. R. Feingold et al., Eds. (South Dartmouth (MA), 2000).

14. K. Sharma et al., Endogenous adenine mediates kidney injury in diabetic models and predicts diabetic kidney disease in patients. J Clin Invest, (2023).

15. T. Suhara, Y. Baba, B. K. Shimada, J. K. Higa, T. Matsui, The mTOR Signaling Pathway in Myocardial Dysfunction in Type 2 Diabetes Mellitus. Curr Diab Rep 17, 38 (2017).

16. M. Chaffin et al., Single-nucleus profiling of human dilated and hypertrophic cardiomyopathy. Nature 608, 174–180 (2022).

17. S. L. Pek et al., Elevation of a novel angiogenic factor, leucine-rich-alpha2-glycoprotein (LRG1), is associated with arterial stiffness, endothelial dysfunction, and peripheral arterial disease in patients with type 2 diabetes. J Clin Endocrinol Metab 100, 1586–1593 (2015).

18. J. J. Liu et al., Association of Plasma Leucine-Rich alpha-2 Glycoprotein 1, a Modulator of Transforming Growth Factor-beta Signaling Pathway, With Incident Heart Failure in Individuals With Type 2 Diabetes. Diabetes Care 44, 571–577 (2021).

19. G. G. Schiattarella et al., Nitrosative stress drives heart failure with preserved ejection fraction. Nature 568, 351–356 (2019).

20. D. An, B. Rodrigues, Role of changes in cardiac metabolism in development of diabetic cardiomyopathy. Am J Physiol Heart Circ Physiol 291, H1489–1506 (2006).

21. L. M. Semeniuk, A. J. Kryski, D. L. Severson, Echocardiographic assessment of cardiac function in diabetic db/db and transgenic db/db-hGLUT4 mice. Am J Physiol Heart Circ Physiol 283, H976–982 (2002).

22. T. Oka et al., Mitochondrial DNA that escapes from autophagy causes inflammation and heart failure. Nature 485, 251–255 (2012).

23. R. Iorio, G. Celenza, S. Petricca, Mitophagy: Molecular Mechanisms, New Concepts on Parkin Activation and the Emerging Role of AMPK/ULK1 Axis. Cells 11, (2021).

24. M. L. Seibenhener et al., Sequestosome 1/p62 is a polyubiquitin chain binding protein involved in ubiquitin proteasome degradation. Mol Cell Biol 24, 8055–8068 (2004).

25. S. Pankiv et al., p62/SQSTM1 binds directly to Atg8/LC3 to facilitate degradation of ubiquitinated protein aggregates by autophagy. J Biol Chem 282, 24131–24145 (2007).

26. G. Ashrafi, T. L. Schwarz, The pathways of mitophagy for quality control and clearance of mitochondria. Cell Death & Differentiation 20, 31–42 (2013).

27. A. S. Strimpakos, E. M. Karapanagiotou, M. W. Saif, K. N. Syrigos, The role of mTOR in the management of solid tumors: an overview. Cancer Treat Rev 35, 148–159 (2009).

28. R. Yuan, A. Kay, W. J. Berg, D. Lebwohl, Targeting tumorigenesis: development and use of mTOR inhibitors in cancer therapy. J Hematol Oncol 2, 45 (2009).

29. J. König et al., Mitochondrial contribution to lipofuscin formation. Redox Biol 11, 673–681 (2017).

30. O. Mgbemena, Y. Zhang, G. Velarde, Role of Diabetes Mellitus in Heart Failure With Preserved Ejection Fraction: A Review Article. Cureus 13, e19398 (2021).

31. A. Pandey, M. S. Khan, K. V. Patel, D. L. Bhatt, S. Verma, Predicting and preventing heart failure in type 2 diabetes. Lancet Diabetes Endocrinol 11, 607–624 (2023).

32. H. Koenig, A. D. Goldstone, C. Y. Lu, Polyamines are intracellular messengers in the beta-adrenergic regulation of Ca2+ fluxes, [Ca2+]i and membrane transport in rat heart myocytes. Biochem Biophys Res Commun 153, 1179–1185 (1988).

33. W. Durante, L. Liao, S. V. Reyna, K. J. Peyton, A. I. Schafer, Transforming growth factor-beta(1) stimulates L-arginine transport and metabolism in vascular smooth muscle cells: role in polyamine and collagen synthesis. Circulation 103, 1121–1127 (2001).

34. H. Li et al., Regulatory role of arginase I and II in nitric oxide, polyamine, and proline syntheses in endothelial cells. Am J Physiol Endocrinol Metab 280, E75–82 (2001).

35. G. Jia, M. A. Hill, J. R. Sowers, Diabetic Cardiomyopathy: An Update of Mechanisms Contributing to This Clinical Entity. Circ Res 122, 624–638 (2018).

36. R. A. Saxton, D. M. Sabatini, mTOR Signaling in Growth, Metabolism, and Disease. Cell 168, 960–976 (2017).

